# Point-of-care cardiac elastography with external vibration for quantification of diastolic myocardial stiffness

**DOI:** 10.1101/2024.01.26.24301851

**Authors:** Tom Meyer, Brunhilde Wellge, Gina Barzen, Fabian Knebel, Katrin Hahn, Thomas Elgeti, Thomas Fischer, Jürgen Braun, Heiko Tzschätzsch, Ingolf Sack

## Abstract

**Background:** Heart failure is an increasing health problem in Western societies. Approximately 50% of patients with heart failure have preserved ejection fraction (HFpEF) and concomitant diastolic dysfunction (DD), in part caused by increased myocardial stiffness not detectable by standard echocardiography. While elastography can map tissue stiffness, cardiac applications are currently limited, especially in patients with a higher body mass index (BMI). Therefore, we developed point-of-care cardiac elastography to detect abnormal diastolic myocardial stiffness associated with DD.

**Methods:** Cardiac time-harmonic elastography (THE) using standard medical ultrasound and continuous external vibration was developed and applied to healthy controls and participants with DD due to wild-type transthyretin amyloidosis (ATTR) in this prospective single-center study between June 2020 and December 2022. A subgroup of participants with ATTR was on tafamidis treatment. Diastolic shear wave speed (SWS) was determined as surrogate marker of myocardial stiffness in different cardiac regions including the septum, posterior wall and automatically detected global left ventricular wall.

**Results:** A total of 130 participants were screened and 44 participants with ATTR (4 women, mean age: 80±7 years, BMI range: 20-37) and 54 healthy controls (26 women, mean age: 47±16 years, BMI range: 15-32) were included. In all analyzed regions, SWS was higher in patients than in controls providing area-under-the-curve (AUC) values (septum: 1.8±0.3m/s versus 2.9±0.6m/s, AUC=0.996; posterior wall: 1.9±0.3m/s versus 2.7±0.5m/s, AUC=0.938; global left ventricular wall: 2.0±0.3m/s versus 2.6±0.4m/s, AUC=0.912). Furthermore, SWS was reduced in participants treated with tafamidis (septum: 2.6±0.4 m/s; posterior wall: 2.4±0.3m/s; global left ventricular wall: 2.3±0.3m/s, all p<.005) suggesting the use of THE for therapy monitoring and patient management.

**Conclusions:** Cardiac THE detects abnormal myocardial stiffness in patients with DD, independent of BMI and ROI selection. Because it uses standard ultrasound components, THE can be cost-effectively implemented as a point-of-care device for widespread clinical use.

## Introduction

Heart failure is a public health concern with increasing prevalence(1, 2) and related hospital admissions placing a significant financial burden on the healthcare systems in Western societies(2). Approximately half of patients with heart failure have a preserved ejection fraction (HFpEF)(3). The main pathophysiological hallmark of HFpEF is diastolic dysfunction (DD)(4), caused in part by abnormally high left ventricular (LV) myocardial stiffness(5).

Because stiffness measurements require the application of mechanical stress, myocardial stiffness cannot be quantified by standard echocardiography(6).

Elastography is a specialized imaging technique for noninvasive measurement of tissue stiffness. Quantitative elastography relies on the measurement of propagating shear waves induced endogenously by cardiac valve closure(7), acoustic radiation force impulses (ARFI)(8), or by external vibrations(9). The shear waves can be captured by standard imaging techniques like ultrasound(10) or magnetic resonance imaging (MRI)(11) and then converted into stiffness maps(7, 12, 13).

Combined with established echocardiography, cardiac elastography could provide a holistic, noninvasive and quantitative picture of the mechanical work of the heart, helping physicians to detect altered cardiac function in the early stages of disease. However, promising approaches currently under clinical evaluation rely on expensive hardware(14), either MRI(12, 15) or multichannel ultrasound(7, 8, 16–22), which hinders their widespread clinical use(23). In addition, MRI-based cardiac elastography is time-consuming, endogenous shear wave elastography is limited to specific cardiac events and regions and ARFI-based methods are limited in depth(23), which is relevant given obesity as a risk factor for HFpEF(24).

Time-harmonic elastography (THE) relies on standard line-by-line ultrasound acquisition and externally induced low-frequency vibrations(9). The low-frequency waves penetrate deep into the body with little attenuation independent of body-mass-index (BMI)(25). THE generates stiffness maps in the whole field-of-view and has shown its effectiveness in several organs including the liver(9), brain(26) and aorta(27). However, THE has never been used for quantitative mapping of myocardial stiffness.

Therefore, we developed and present here cardiac THE for the quantification of myocardial stiffness. The clinical utility of cardiac THE is demonstrated in participants with wild-type transthyretin amyloidosis (ATTR), which is associated with increased left ventricular (LV) wall thickness and diastolic myocardial stiffness(28) and for which drug treatment is available(29).

Our aim is to provide a new echocardiographic method that i) quantifies stiffness throughout the LV at greater depths, ii) is easy to use with little operator interaction for regional analysis, iii) is cost-effective and can be distributed to cardiology centers to improve diagnosis and management of cardiac disease associated with abnormal myocardial stiffening.

## Material and Methods

### Subjects

The study was approved by the institutional review board (EA1/086/15). Written informed consent for study participation was obtained from all subjects before inclusion. A total of 130 participants were prospectively screened for enrolment from June 2020 to December 2022. 44 participants with verified wild-type ATTR amyloidosis according to the criteria of the European Society of Cardiology(30) and confirmed by bone scintigraphy and genetic tests to rule out hereditary ATTR, were included in this study.

54 participants without known major cardiac disease and no DD (age-adapted) according to the echocardiography guidelines(5), were used as healthy controls. A subgroup of 11 participants with ATTR was on treatment with tafamidis (61mg once daily; Pfizer, New York, USA) for a duration of 14±10 (range: 2-36) months.

Participants with a history of myocardial infarction or stroke within 2 weeks before the investigation, unstable angina pectoris, implantable cardioverter-defibrillators, poor echogenicity, or inability to follow breathing instructions were excluded from our analysis. The recruitment scheme is shown in figure 1. Demographic data are summarized in table 1. All participants underwent THE and a full echocardiographic examination. 10 healthy participants were re-investigated after 2-6 months to analyze test-retest reproducibility.

**Figure 1:**
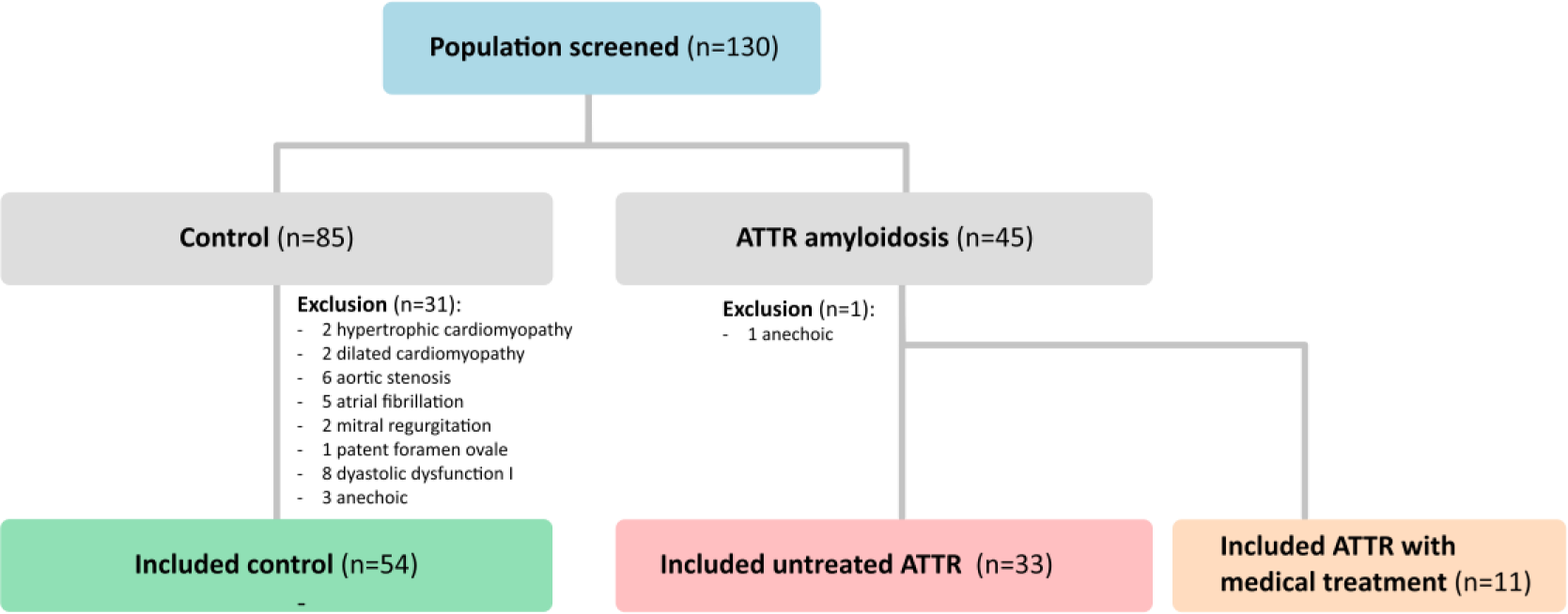
130 subjects were screened for inclusion. The analysis included 54 healthy controls and 44 participants with wild-type amyloid transthyretin (ATTR), of whom 11 were on medical treatment with tafamidis.

**Table 1:** Demography, echocardiography, and cardiac THE values by group reported as mean±standard deviation along with *p*-values for group differences. Ranges are reported in squared brackets.

|  | Mean ± SD |  |  | *p*-values |  |  |
| --- | --- | --- | --- | --- | --- | --- |
|  | CTR<br>(n = 54) | Untreated<br>ATTR<br>(n = 33) | treated ATTR<br>(n = 11) | CTR vs<br>untreated<br>ATTR | CTR vs<br>treated ATTR | untreated vs<br>treated ATTR |
| **Demography** |  |  |  |  |  |  |
| Age (years) | 47 ± 16<br>[24-79] | 80 ± 7<br>[53-91] | 79 ± 9<br>[66-90] | <0.001 | <0.001 | .76 |
| Sex (f/m) | 26 / 28 | 4 / 29 | 0 / 11 | n.a. | n.a. | n.a. |
| BMI (kg/m<sup>2</sup>) | 24 ± 4<br>[15-32] | 25 ± 4<br>[20-37] | 26 ± 4<br>[21-33] | .1 | .23 | .93 |
| Heart rate (bpm) | 68 ± 11 | 77 ± 17 | 73 ± 10 | .01 | .18 | .34 |
| **Echocardiography** |  |  |  |  |  |  |
| IVSd (mm) | 9 ± 2 | 19 ± 4 | 20 ± 4 | <.001 | <.001 | .2 |
| LVPWd (mm) | 9 ± 1 | 16 ± 4 | 16 ± 4 | <.001 | <.001 | .97 |
| LAVI | 29 ± 7 | 51 ± 15 | 49 ± 13 | <.001 | <.001 | .64 |
| GLS (%) | 19 ± 3 | 11 ± 3 | 13 ± 3 | <.001 | <.001 | .046 |
| EF (%) | 60 ± 5 | 48 ± 8 | 51 ± 7 | <.001 | .001 | .16 |
| E (m/s) | 0.8 ± 0.2 | 0.9 ± 0.2 | 1 ± 0.3 | .1 | .03 | .12 |
| E/e' | 7 ± 3 | 18 ± 7 | 21 ± 6 | <.001 | <.001 | .18 |
| e' (cm/s) | 12 ± 4 | 5 ± 2 | 5 ± 2 | <.001 | <.001 | .69 |
| E/A* | 1.3 ± 0.5 | 2 ± 1 | 2.2 ± 1 | .01 | .1 | .71 |
| **Cardiac THE** |  |  |  |  |  |  |
| SWS (septum) (m/s) | 1.8 ± 0.3 | 2.9 ± 0.6 | 2.6 ± 0.4 | <.001 | <.001 | .04 |
| SWS (post. wall) (m/s) | 1.9 ± 0.3 | 2.7 ± 0.5 | 2.4 ± 0.4 | <.001 | .007 | .09 |
| SWS (global) (m/s) | 2 ± 0.3 | 2.6 ± 0.4 | 2.4 ± 0.3 | <.001 | .003 | .04 |
| Depth (septum) (cm) | 6 ± 1 | 7 ± 1 | 7 ± 1 | <.001 | <.001 | .51 |
| Depth (post. wall) (cm) | 11 ± 2 | 12 ± 1 | 12 ± 1 | <.001 | <.001 | .61 |
| Amplitude (septum) (μm) | 0.72 ± 0.38 | 0.92 ± 0.76 | 0.71 ± 0.45 | .16 | .96 | .27 |
| Amplitude (post. wall) (μm) | 0.68 ± 0.41 | 1.07 ± 0.77 | 0.94 ± 0.71 | .01 | .26 | .62 |

\* could not be obtained in 24 patients due to atrial fibrillation
BMI=body mass index; IVSd=Interventricular septum thickness in diastole; LVPWd=left ventricular posterior wall thickness in diastole; LAVI=left atrial volume index; GLS=global longitudinal strain; EF=ejection fraction; e'=early diastolic mitral annular velocity; E=early diastolic peak; A=late diastolic peak; SWS=shear wave speed

### Echocardiography

The subjects were positioned on a vibration bed (GAMPT, Merseburg, Germany) as required for the subsequent THE examination. A trained cardiologist (BW) performed echocardiography according to recent recommendations(31, 32) using a Vivid E9 scanner (GE Healthcare, Horton, Norway) with an M5Sc transducer.

### Time Harmonic Elastography (THE)

THE utilized a vibrating bed, as shown in Figure 2, to introduce shear waves into the heart and create a mechanical stiffness contrast(9). The waves, stimulated by a superposition of 60 and 70 Hz continuous vibration frequency, were acquired and processed using a clinical ultrasound scanner equipped with a phased array transducer of 2 MHz center frequency (GAMPT, Merseburg, Germany). THE was performed directly after echocardiography in the left lateral decubitus position with the vibration unit positioned below the heart to ensure sufficiently high wave amplitudes. Images were acquired over 2 s at end-expiration in parasternal long-axis view with a depth of up to 15 cm. The acquisitions were not synchronized to the ECG; however, the ECG signal was recorded to automatically identify the diastolic cardiac phases during post-processing, as described in the Supplementary Information.

**Figure 2:**
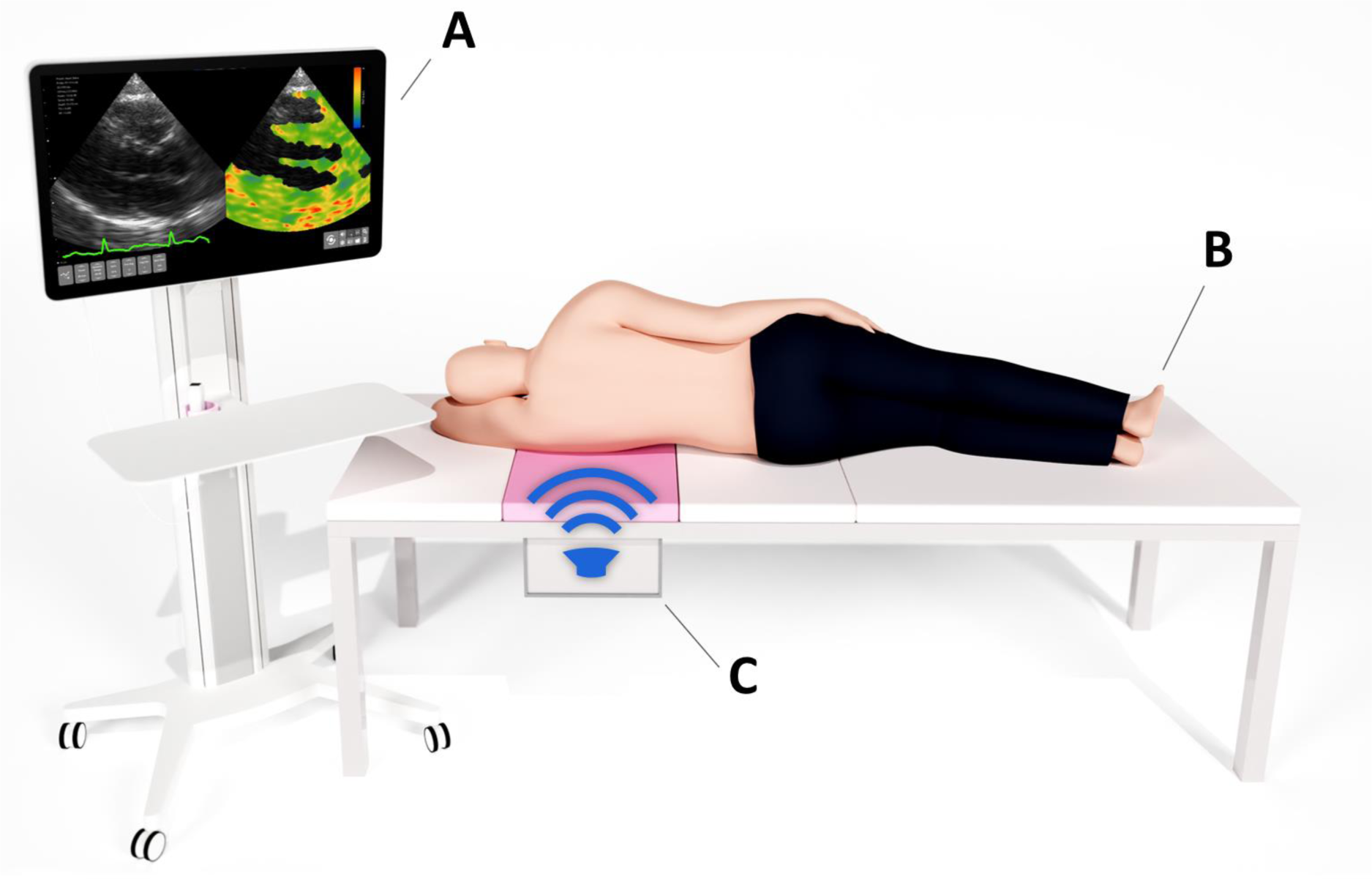
Cardiac time-harmonic elastography (THE) setup. (A) Customized clinical ultrasound scanner for data acquisition and data processing. (B) Vibration bed with integrated vibration unit (pink, C) for induction of harmonic multifrequency vibrations.

Measurements were repeated 5 times to average the final diastolic stiffness maps generated by the post-processing algorithm (13), which is described in detail in the supplemental material S1. In brief: First, the axial displacement between acquired ultrasound frames was computed based on the phase difference of adaptive windows following the cardiac wall motion. Then, spatio-temporal noise was suppressed using singular value decomposition for denoising. Next, vibration frequencies were isolated using Fourier-based band pass filters for both fundamental vibration frequencies. Finally, the obtained time- and frequency resolved wavefields were converted into shear wave speed (SWS in m/s) maps using wave-number based k-MDEV gradient inversion(13), which quantifies the local phase-gradient of the waves. Diastolic SWS maps were averaged and displayed as surrogate parameter for diastolic stiffness.

Three regions of interest (ROI) were analyzed, as indicated in figure 3: Two ROI, manually drawn by one observer, covering (i) the midsection of the septum, and (ii) the posterior wall excluding the pericardium. Additionally, one ROI included automatically identified myocardium based on a region-growing algorithm with adaptive thresholding of B-mode images (iii). SWS values were averaged within each ROI. Medians of repeated measurements were tabulated as diastolic SWS values (see table 1).

**Figure 3:**
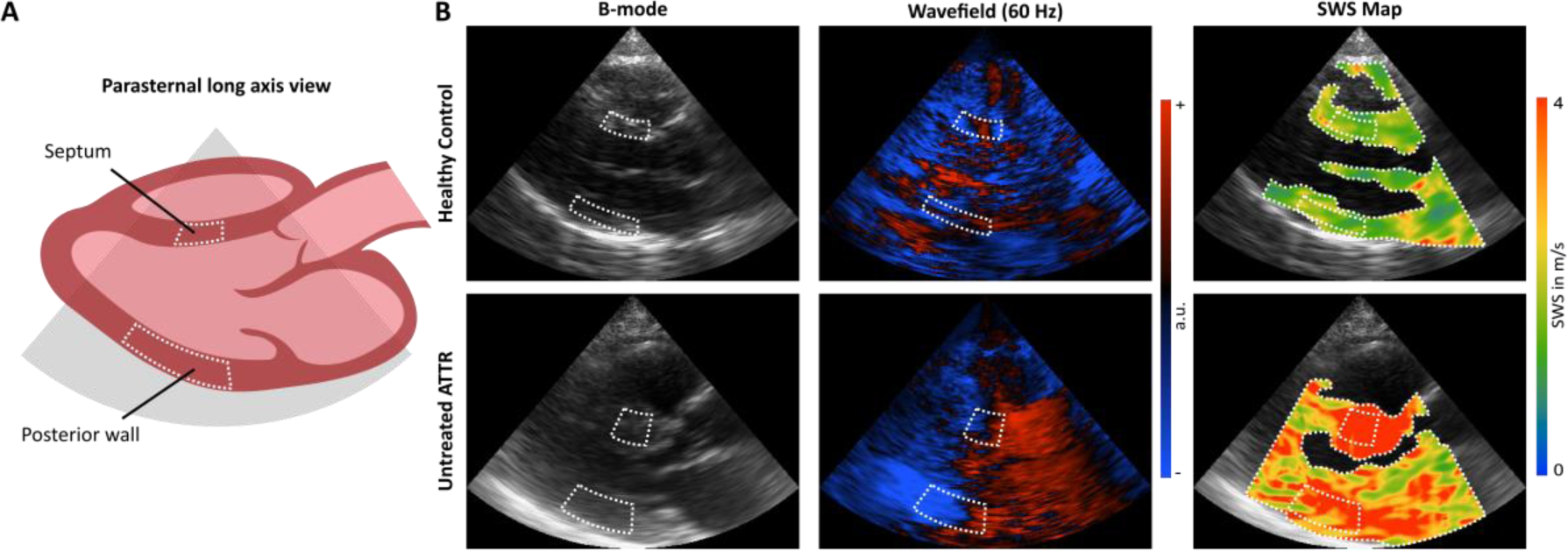
(A) Anatomical overview of the measurements performed in the parasternal long-axis view. (B) Representative B-mode images and wavefields at 60Hz and shear wave speed (SWS) maps of a healthy control subject and an untreated wild-type amyloid-transthyretin (ATTR) participant. Regions of interest (ROI) are demarcated by white dotted lines. ROI of the septum and posterior wall were manually selected. A third (global) ROI was automatically identified to select a larger region covering the major portions of the myocardium visible in the image.

### Statistics

Differences between regions and groups were tested using Student’s t-test. Correlations between SWS, demographic-, and echocardiographic parameters were tested using Pearson’s correlation coefficient. The influence of age was analyzed using linear regression. The discriminative power of SWS was estimated using receiver-operator characteristic (ROC) curve analysis. 95% confidence intervals were obtained via bootstrapping. Youden’s index was calculated to identify optimal SWS cutoffs in each region. Corresponding sensitivity and specificity are reported. Bonferroni correction was used to correct for multiple testing. Test-retest reproducibility was analyzed using the one-way random intraclass correlation coefficient (ICC). All values were tabulated as mean±SD with ranges in square brackets, unless otherwise stated. Corrected p-values <.05 were considered significant. The group size was estimated based on a power analysis assuming effect sizes of 0.6 to 0.7 at a 5% significance level and a power of 0.8 according to the reported variability of stiffness parameters encountered in healthy adult and ATTR patients(8, 15, 17, 19). Statistical analysis was performed in R (version 4.1.1;R-Foundation,Vienna,Austria) using R-Studio (version 2021.09.0;RStudio,PBC,Boston,MA).

## Results

Demographic, echocardiographic, and THE results are summarized in table 1. Mean age was lower in controls (47±16 [24-79] years) than in participants with ATTR (80±7 [53-91] years), motivating a subgroup analysis of SWS within overlapping age ranges (controls: n=19, age: 64±8 [53-79] years, ATTR: n=13, age: 73±7 [53-79] years), which is summarized in table 2. While BMI was not different between controls (24±4 [15-32]) and participants with untreated ATTR (25±4 [20-37]), the latter had significantly altered heart morphology, as evidenced by greater myocardial wall thickness (p=0.003, see table 1). As expected, echocardiographic parameters describing LV function were significantly different between controls and participants with ATTR (see table 1). There were no differences in demographic or echocardiographic parameters between untreated and treated participants.

**Table 2:** Age-matched subgroup analysis: Demography, echocardiography, and cardiac elastography THE values by group reported as mean±standard deviation along with *p*-values for age matched group differences. Ranges are reported in squared brackets.

|  | Mean ± SD<br>Age Matched |  | *p*-values<br>Age Matched |
| --- | --- | --- | --- |
|  | CTR<br>(n = 19) | untreated ATTR<br>(n = 13) | CTR vs untreated<br>ATTR |
| **Demography** |  |  |  |
| Age (years) | 64 ± 8<br>[53-79] | 73 ± 7<br>[53-79] | .001 |
| Sex (f/m) | 7 / 12 | 1 / 12 | n.a. |
| BMI (kg/m<sup>2</sup>) | 23 ± 4<br>[15-29] | 26 ± 5<br>[20-37] | .1 |
| Heart rate (bpm) | 68 ± 10 | 81 ± 19 | .03 |
| **Echocardiography** |  |  |  |
| IVSd (mm) | 10 ± 2 | 18 ± 3 | <.001 |
| LVPWd (mm) | 10 ± 2 | 17 ± 3 | <.001 |
| LAVI | 30 ± 8 | 46 ± 10 | <.001 |
| GLS (%) | 19 ± 3 | 10 ± 3 | <.001 |
| EF (%) | 61 ± 6 | 46 ± 9 | .07 |
| E (m/s) | 0,7 ± 0,2 | 0.8 ± 0.3 | .25 |
| E/e' | 8 ± 4 | 15 ± 4 | <.001 |
| e' (cm/s) | 10 ± 3 | 6 ± 2 | <.001 |
| E/A* | 1 ± 0,5 | 1.9 ± 1.2 | .07 |
| **Cardiac THE** |  |  |  |
| SWS (septum) (m/s) | 1,9 ± 0,3 | 2.8 ± 0.5 | <.001 |
| SWS (post. wall) (m/s) | 2 ± 0,3 | 2.5 ± 0.4 | <.001 |
| SWS (global) (m/s) | 2,1 ± 0,3 | 2.4 ± 0.3 | .007 |
| Depth (septum) (cm) | 7 ± 1 | 7 ± 1 | .08 |
| Depth (post. wall) (cm) | 11 ± 1 | 12 ± 1 | .009 |
| Amplitude (septum) (μm) | 0,68 ± 0,32 | 0.68 ± 0.28 | 1.0 |
| Amplitude (post. wall) (μm) | 0,71 ± 0,5 | 0.78 ± 0.44 | .66 |

\* could not be obtained in 24 participants due to atrial fibrillation
BMI=body mass index; IVSd=Interventricular septum thickness in diastole; LVPWd=left ventricular posterior wall thickness in diastole; LAVI=left atrial volume index; GLS=global longitudinal strain; EF=ejection fraction; e'=early diastolic mitral annular velocity; E=early diastolic peak; A=late diastolic peak; SWS=shear wave speed

THE provided SWS maps of diastolic myocardium (figure 3) for all subjects. Group comparisons are shown in figure 4. In controls, SWS was 1.8±0.3m/s in the septum, 1.9±0.3m/s in the posterior wall, and 2.0±0.3m/s globally. Excellent reproducibility was found with ICC=0.82, 0.82 and 0.79 for septum, posterior wall and global ROI, respectively. SWS varied slightly between regions, with lower values in the septum than in the posterior wall and globally (septum vs. posterior wall: p<.001; septum vs. global: p<.001; posterior wall vs. global: p=.036), indicating a rather homogeneous distribution of myocardial stiffness, especially in untreated ATTR participants, where regional differences were found only between the septum and global ROIs (p=.004).

**Figure 4:**
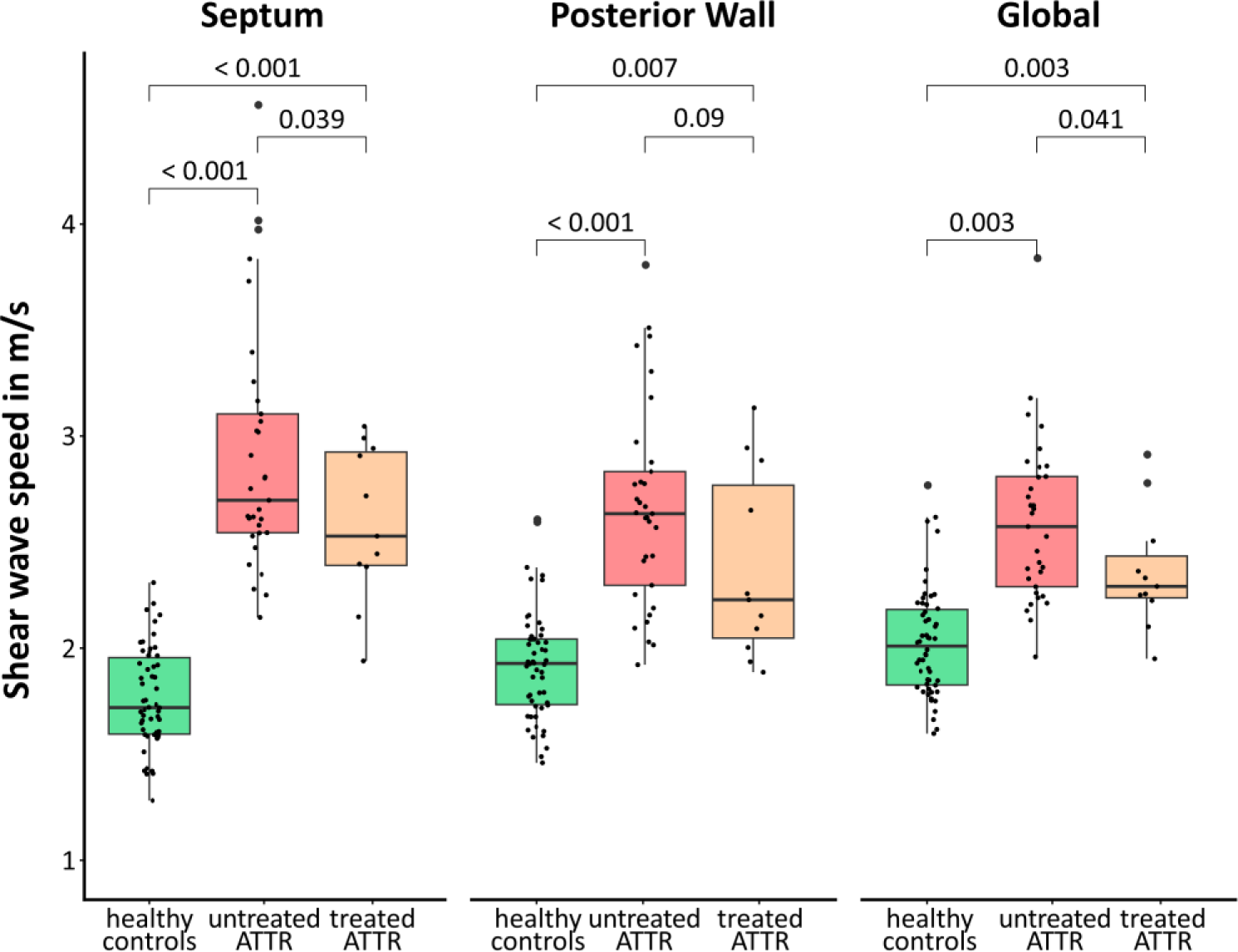
Shear wave speed of the septum, posterior wall and the automatically masked, global myocardium in healthy controls, untreated wild-type amyloid transthyretin (ATTR) patients, and treated ATTR patients. Stiffness is elevated in all regions in patients compared to controls.

Compared to controls, participants with untreated ATTR showed 1.6-, 1.4- and 1.3-fold higher SWS with excellent discriminative powers (AUC=0.996, 0.938, 0.912, respectively) for the three regions (see figure 5). Subgroup analysis of SWS within overlapping age ranges also revealed high AUC values for myocardial stiffening due to ATTR in all three regions (0.991, 0.866, 0.793). Participants treated with tafamidis had slightly lower SWS values (septum: p=.039; global region: p=.041) than untreated participants.

**Figure 5:**
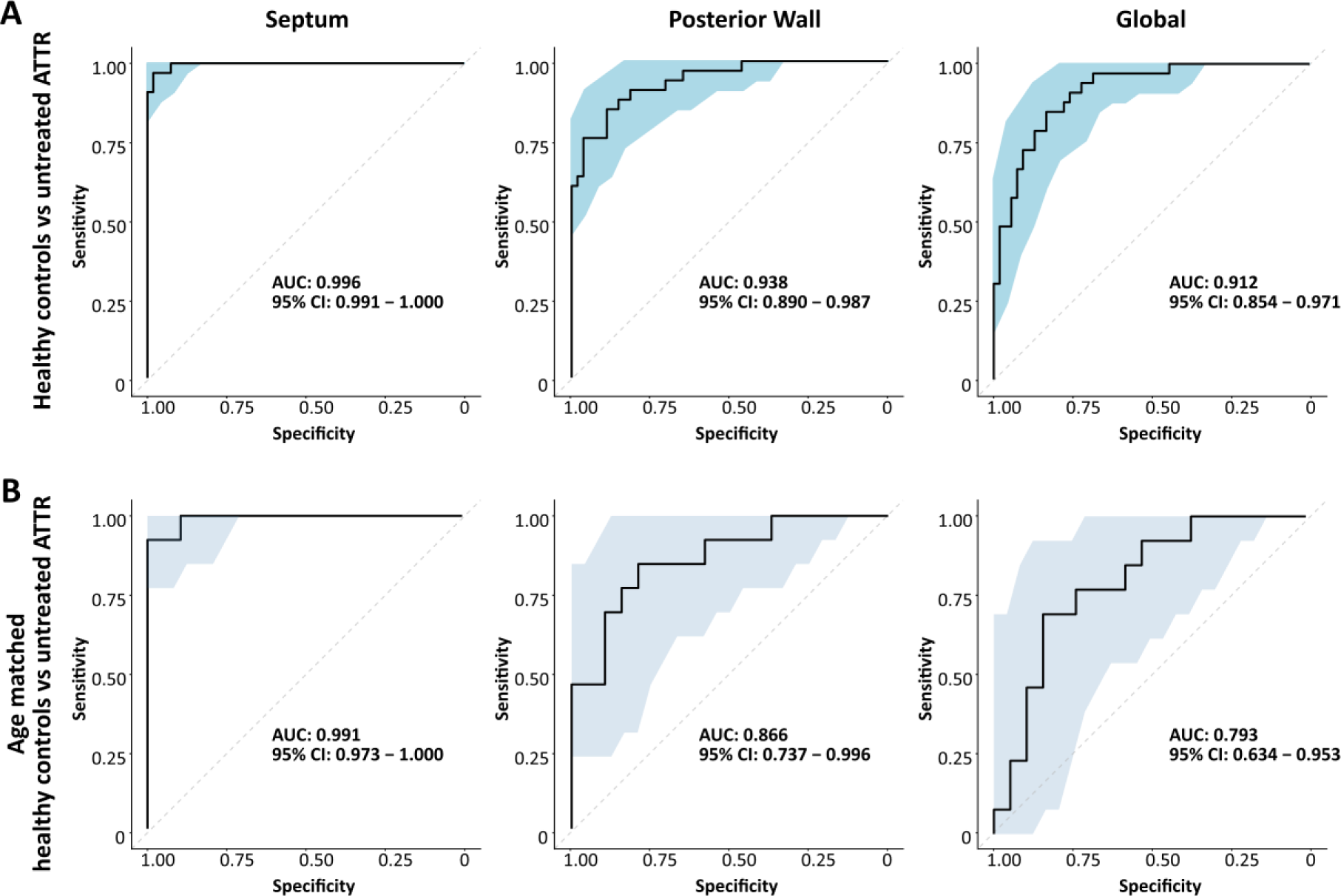
Receiver operator characteristic curves for the discrimination of healthy controls from wild-type amyloid transthyretin (ATTR) patients (A), age matched ATTR patients (B) based on myocardial stiffness in the septum, posterior wall, and globally averaged over the left ventricle, measured by cardiac time-harmonic elastography.

Correlations between SWS and echocardiographic and demographic parameters were analyzed separately for each group. In the analysis of age effects (figure 6A), a moderate correlation was found only for SWS of the septum (R=0.33, p=.015). Linear regression analysis yielded an SWS increase of 0.005m/s per year. For participants with ATTR, global SWS correlated with age (R=0.35, p=.04) in the treated group, and SWS in the septum correlated negatively with age (R=-0.68, p=.02) in the untreated group. Most echocardiographic parameters showed no correlation with SWS. In particular, no correlation was found between SWS and septal or posterior wall thickness (figure 6B). However, negative correlations were found between global longitudinal strain (GLS) and SWS in the posterior wall (R=-0.51, p=.004) and global region (R=-0.41, p=.003) in controls, and global SWS correlated with left atrial volume index (LAVI; R=0.51, p=.002) in participants with untreated ATTR. No correlation was found between BMI and SWS in any group or subgroup.

**Figure 6:**
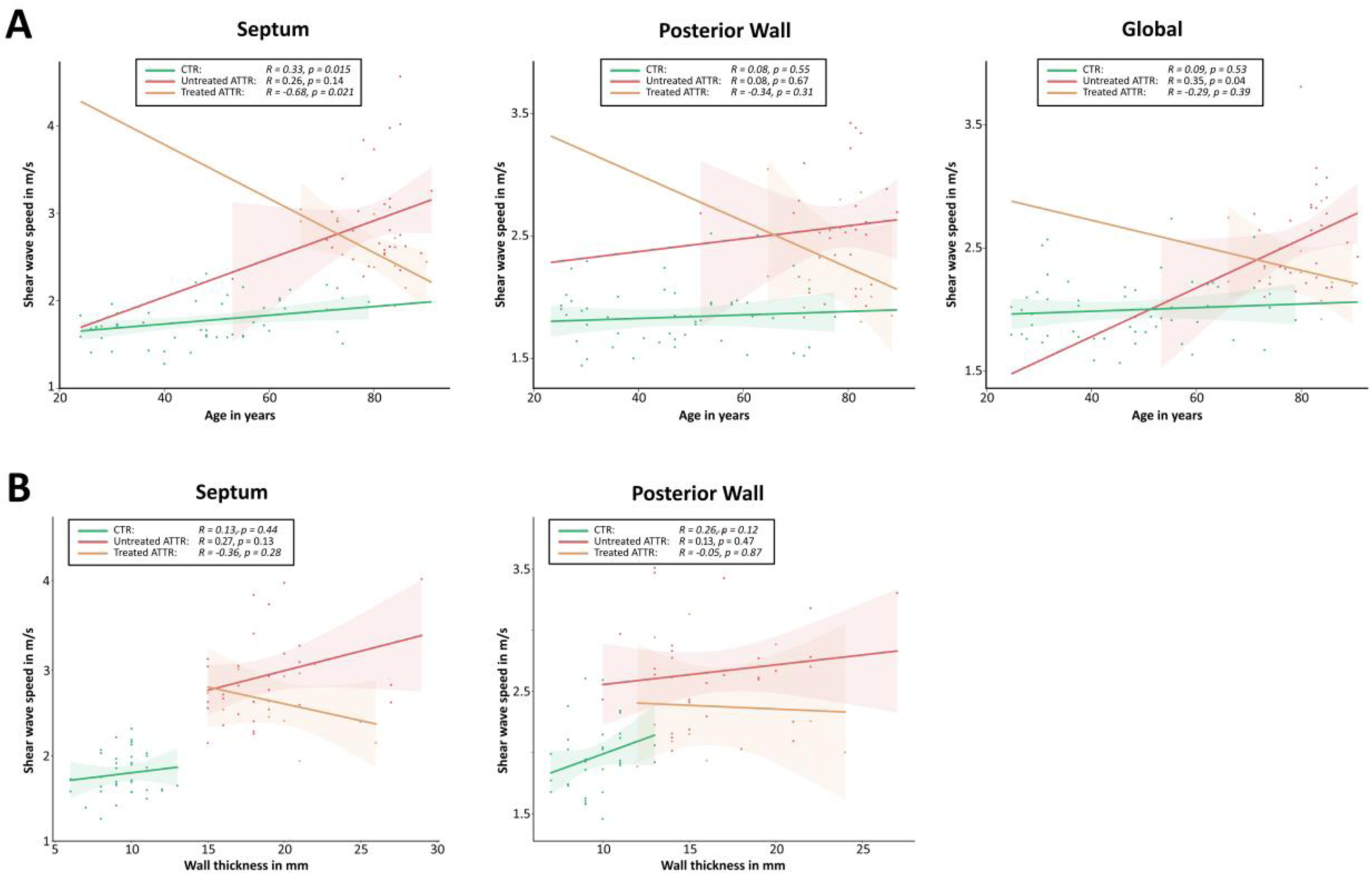
Scatterplots showing the correlation of each age (A) and wall thickness (B) with shear wave speed (SWS) in healthy controls (CTR) and ATTR patients with wild-type transthyretin amyloidosis (ATTR) untreated and treated with tafamidis medication.

## Discussion

To the best of our knowledge, this is the first study to quantify in vivo myocardial stiffness in adults by ultrasound elastography with external harmonic vibration. Using cardiac THE with standard line-by-line acquisition, implemented in a point-of-care device without the need for expensive multichannel hardware, we were able to generate quantitative maps of diastolic myocardial stiffness of the septum and posterior wall. Our results demonstrate excellent diagnostic accuracy of cardiac THE in detecting abnormal myocardial stiffness as a hallmark of DD in ATTR patients and its potential clinical use for monitoring treatment responses.

THE provided SWS maps of the full field-of-view with 15cm depth, which resolves several issues with existing methods. Myocardial stiffness could be quantified in several regions based on a single scan, which improves cross-sectional consistency of stiffness data. This includes the posterior wall as an important region for full myocardial assessment(33). Despite obesity (BMI≥30) of eight subjects, all participants could be examined, including the posterior myocardial wall. However, the mean central septal depth in our study population was 6±1cm, exceeding the ARFI-SWEI limit of 7cm(21) in 36 participants.

Several earlier studies investigated diastolic myocardial stiffness using different elastography methods. For comparison, we converted stiffness reported as shear modulus *G*^∗^ to SWS (SWS=√|*G*^∗^|/*ρ*, with unit mass density ρ=1kg/L)(10). Our stiffness values in controls agree with results from ARFI-SWEI, which range from 1.45±0.14m/s(22) to 2.1±0.4m/s(21), and the range reported with MRI elastography at 80Hz of 1.4-2.4m/s(34). As expected, higher SWS values were observed in MRI elastography at higher vibration frequency (140Hz) (2.9[2.7-3.4]m/s)(15). Values similar to ours were also found in studies exploiting the velocity of the myocardial stretch wave after atrial contraction (1.6±0.2m/s)(19). However, many studies based on endogenous waves after mitral valve closure measuring end-diastolic stiffness, including MRI-measured SWS, found higher SWS ranging from 3.2m/s to 4.7m/s(12, 16–18, 20), suggesting myocardial stiffening already in the early phase of isovolumetric contraction.

In agreement with Villemain et al.(21), we observed age-dependent myocardial stiffening in the septum. However, this age effect was less pronounced in our study and similar to the findings of other investigators, who report minor(17) or no(34) age effects, possibly masked by a greater biological variability in our study population, as reflected by a wide BMI-range.

In echocardiography, ATTR cardiomyopathy is usually identified by DD grade 2 or worse, increased wall thickness, and reduced GLS in basal LV regions (“apical sparing”). A concomitant increase in myocardial stiffness due to deposition of transthyretin amyloid fibrils was expected(28) and has been confirmed by our study. Previous studies also observed ATTR-associated myocardial stiffening by factors of 1.2 to 2.0(15, 17, 19). Our AUC values (0.912-0.996) suggest an excellent discriminative power for ATTR versus healthy controls, especially in the septal region. Similar results were reported for different cardiomyopathies associated with wall thickening (AUC=0.99)(21). However, our correlation analysis of wall thickness and myocardial stiffness demonstrates that there is no linear relationship between the two parameters indicating that stiffness alone is an important diagnostic parameter.

Furthermore, our results in the subgroup of participants with ATTR treated with tafamidis suggest that myocardial stiffness reflects drug efficacy as softer values were found in the septum compared to untreated participants. This observation encourages studies with more participants before and during therapy. Since medications in HFpEF are becoming available(29) and life-style interventions have shown high efficacy(35), monitoring myocardial stiffness may support the introduction and validation of novel therapies.

Despite the encouraging results, our study has limitations. First, by nature of its design as a single-center exploratory study, group sizes were too small to draw conclusions about the effect of treatment duration on myocardial stiffness. As the prevalence of ATTR amyloidosis is higher in the elderly, the groups of participants with ATTR and healthy volunteers with overlapping age ranges were small. Technical limitations are current restrictions to 2D acquisition and assessment of diastolic phases only. A possible 2D bias was minimized by spatiotemporal filters, which effectively suppressed waves that cross the image plane obliquely and by the low impact of this artifact at low crossing angles(9). Assessment of systolic phases is currently under development using further optimized displacement estimation for suppressing rapid wall motion. Finally, our current THE setup relies on a vibrating bed not easily moveable to the patient. To further improve the clinical utility of THE, portable actuators are being currently tested that may replace the vibration bed in the future.

In summary, we have developed cardiac THE – an imaging modality based on external vibration and cost-effective, line-by-line ultrasound - for noninvasive quantification of myocardial stiffness. Our results in ATTR patients with and without treatment demonstrate the excellent diagnostic accuracy of THE in detecting abnormal diastolic myocardial stiffness. Incorporated into clinical routine, THE could complement standard echocardiography for improved diagnosis, treatment monitoring, and management of patients with a wide range of diseases involving elevated diastolic myocardial stiffness.

## Data Availability

SWS data generated or analyzed during the study are available from the corresponding author upon reasonable request

## Supplemental Material

### S1. Technical aspects of cardiac time harmonic elastography

Time harmonic elastography (THE) is based on externally induced harmonic vibrations captured with standard line-by-line clinical ultrasound acquisition at relatively low frame rates (1). THE has been validated in phantoms (1, 2) and established for several clinical applications such as the detection of liver fibrosis (3), chronic kidney disease (4), intracranial hypertension (5), or the effects of age and hypertension on aortic stiffness (6). Other concepts in ultrasound elastography such as reverberant shear wave elastography (7, 8) and vibrational shear wave elastography (9) also utilize externally induced harmonic vibrations. In the following, the key technical aspects of cardiac THE acquisition and data processing are briefly summarized.

##### Time harmonic elastography (THE) in a nutshell

THE utilizes externally induced vibrations at relatively low frequencies between 20 and 100 Hz, to avoid strong attenuation and facilitate wave propagation deep into the body (1, 10). These vibrations are continuously applied over several seconds, to ensure a steady flux of shear wave energy (3). The resulting steady-state, time-harmonic tissue deflection can be efficiently sampled at ultrasound frame rates less than the period of the motion (11). This low-frame-rate stroboscopic sampling allows THE to be implemented in low-cost standard ultrasound hardware. In post-processing, local tissue motion is obtained by phase-based displacement estimation (12). The detected displacement contains a mixture of intrinsic motion and externally induced displacement. Therefore, frequency decomposition by frequency band-pass filtering is used to extract the induced harmonic vibration components (6, 13). The resulting time series of complex-valued shear wave images are then processed by wave number inversion to generate time series of shear wave speed maps (14).

### Data acquisition

The key elements of THE are the stimulation of the body with gentle vibrations and the acquisition of images based on standard clinical ultrasound. THE vibration frequencies are typically below 120 Hz, which minimizes attenuation and ensures deep penetration of the shear waves into the body. Continuous wave excitation over 1 to 2 seconds produces a steady state of propagating shear waves that can be visualized by medical ultrasound for probing tissue stiffness. Standard line-by-line ultrasound achieves frame rates on the order of 80 to 100 Hz, which causes stroboscopic undersampling of harmonic motion and aliasing after Fourier transformation. However, since the vibration frequencies and the scanner frame rate are known exactly, the wrapped frequencies can be processed further. This principle of controlled aliasing is further explained in Figure S1_1 (1,10). Although stroboscopic under-sampling is one of the core principles of THE, it could also be implemented in parallel imaging systems (11), which allow higher frame rates in the range of 1 kHz and direct sampling of unwrapped frequencies. However, such implementations come at the cost of more complex and more expensive hardware.

**Figure S1_1:**
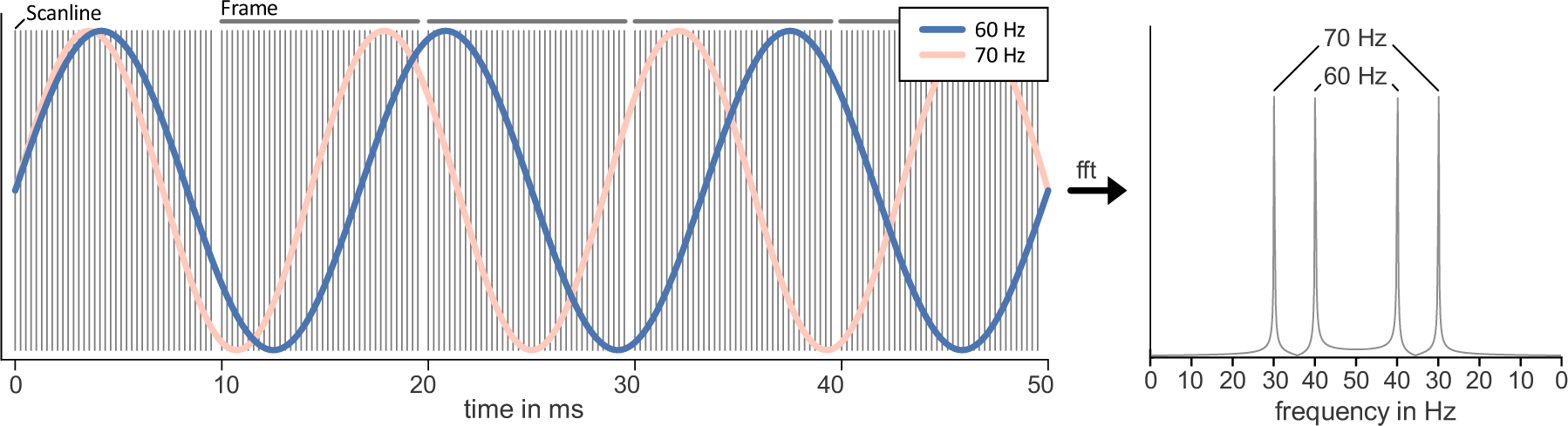
Acquisition sequence for cardiac THE: Each image, acquired at a frame rate of 100 Hz, consisted of 38 scan lines. Multifrequency harmonic vibration at 60 and 70 Hz is under sampled, causing the frequencies to wrap around the Nyquist frequency to 40 and 30 Hz respectively.

In cardiac applications, THE acquires wave data with a frame rate of 100 Hz over two seconds. The increased frame rate as compared to abdominal applications (80 Hz) was chosen to improve the separation of cardiac motion from the desired harmonic vibration signal during post-processing while an acquisition time of two seconds allowed us to cover more than one cardiac cycle. We avoided any further prolongation of the acquisition time because some patients already suffered from breath-hold periods of 2 to 3 seconds. Measurements were performed in a parasternal long axis view with a maximum view depth of 15 cm to simultaneously assess stiffness values in the left ventricular posterior wall and the septal region. It should be noted that although THE is not limited to a specific image window and view angle, the tradeoff between view depth, lateral window size, and frame rate must be carefully balanced.

### Postprocessing

THE processing can be divided into three main steps: (i) estimation of tissue displacements that occurred between acquired ultrasound frames, (ii) generation of frequency-resolved wavefields and (iii) computation of shear wave speed maps using gradient inversion. Each of these steps had to be optimized specifically for cardiac applications. Hyperparameter optimization was performed on a subset of 16 healthy subjects and 16 patients with ATTR. This set of data provided a sufficiently large range of representative cases that ensured optimized post-processing steps from displacement estimation, to frequency decomposition to the reconstruction of shear wave speed maps.

#### 1. Displacement Estimation

Axial displacement between ultrasound frames was estimated from the demodulated, beam-formed, line-by-line radiofrequency data. This was done using Kasai’s algorithm, which is based on the computation of window-wise phase differences between time points (12). In brief, the spatial offset between adjacent time points is obtained by correlation of small windows typically around 1-5 mm in size. This yields both the phase difference, which is translated into axial displacement, and a correlation coefficient, which was used as a quality metric for the estimated displacement.

The wrap limit of this algorithm is determined by the ultrasound wavelength λ/2, which is 380 μm, considering adjacent frames in cardiac THE at 2-MHz center frequency. However, this limit often does not permit reliable estimation of small time-harmonic deflections superimposed on large intrinsic tissue motion. Therefore, displacement estimation was further optimized to account for large tissue motion, such as that present in the heart. As a first step, we computed the displacement frame by frame, using the median filtered estimate of the previous frame as an initial guess for the position of the search window of the next frame. By using the displacement information of the previous frame as a starting point, wraps only occur if the change in velocity exceeds the wrap limit. As a second step, we used singular value decomposition (SVD)-based denoising to remove residual spatiotemporal noise (see Figure S1_2).

It should be noted that THE captures the projection of waves propagating in all directions within the image plane, although it is currently limited to axial displacement estimation. However, adding lateral displacements to shear wave inversion could potentially further improve THE in the future.

**Figure S1_2:**
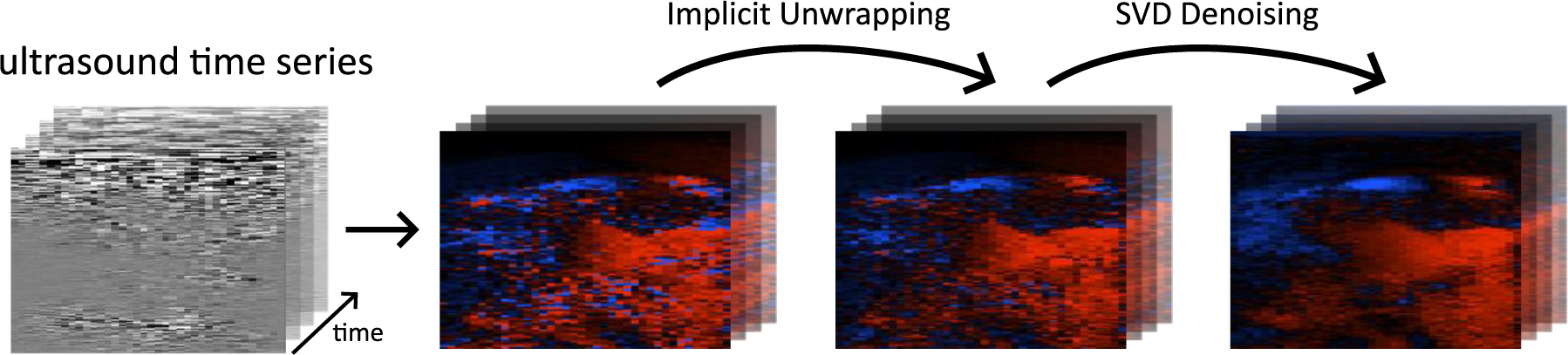
Main steps of displacement estimation for cardiac THE: Displacement is estimated using Kasai’s algorithm(12). Implicit unwrapping stabilizes the estimation with respect to phase wraps while SVD based denoising suppresses spatio-temporal noise.

##### Reconstruction of Wavefields

The resulting displacement fields contain intrinsic cardiac motion and harmonic multifrequency vibrations. The harmonic vibrations were separated from cardiac motion using Hilbert transform (1, 11). Unlike static THE, where the frequency selection is narrow-band, cardiac THE is based on a wider band of frequencies to allow for time resolution. The width of the frequency band determines the temporal resolution after Hilbert transformation. Similar to THE of the aortic wall and the brain and magnetic resonance elastography of the brain (6, 13, 15), we used Butterworth bandpass filters of order 5 and width ±3 Hz (see Figure S1_3).

**Figure S1_3:**
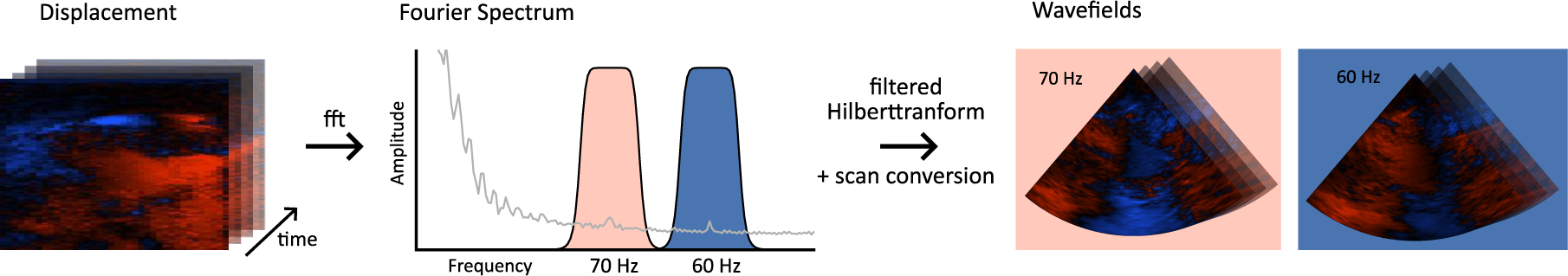
Generation of time- and frequency resolved wavefields: Frequencies were selected using Butterworth bandpass filters in the frequency domain and time-resolved wavefields for each frequency are obtained using the Hilbert transform. Finally, the wavefields are scan-converted to fit the transducer geometry.

#### 2. Gradient Inversion

Time- and frequency-resolved wavefields were converted to shear wave speed (SWS) maps using wave number based *k*-MDEV gradient inversion (1, 14). In simple terms, k-MDEV analyses the local phase-gradient of plane shear waves using first-order 2D derivatives. In mathematical notation, k-MDEV assumes plane shear waves *u*(*r*, *t*) with complex wave number *k*^∗^ (*k*^∗^ = *k*^′^ + *ik*′′, where *k’* and *k’’* denote the real and imaginary parts of *k\**, respectively). With *ω*, the angular frequency, and *u*_0_, the real-valued wave amplitude, the plane wave equation is given by:

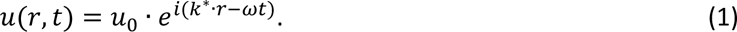

Each complex-valued wave images *u*(*r*, *t*) was decomposed into multiple wave images *ũ*_*df*_ for eight propagation directions (*d*) and two frequencies (*f*) using directional bandpass filters (see Figure S1_4) (16).

**Figure S1_4:**
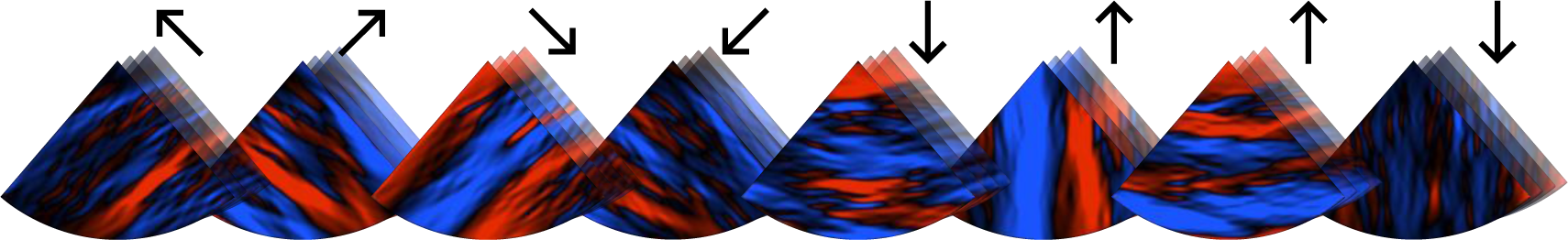
Representative wave field decomposed into 8 plane shear waves for each propagation direction (indicated by the arrows).

Additionally, these bandpass filters suppressed noise and compression wave components. Wave numbers were then derived from the phase gradient of the wave fields *ũ*_*df*_:

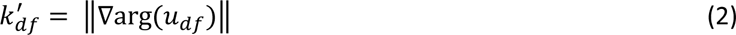

SWS maps were obtained by amplitude-weighted averaging and inversion of *k*^′^

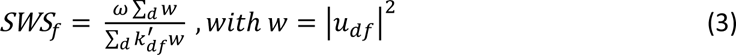

and averaging the inverse *SWS* over frequencies:

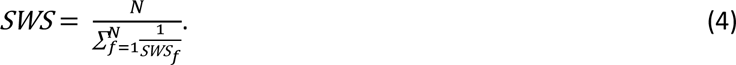

*SWS* is related to the complex shear modulus *G*^∗^ with its real part (storage modulus) G’ and its imaginary part (loss modulus) G’’ as follows (17):

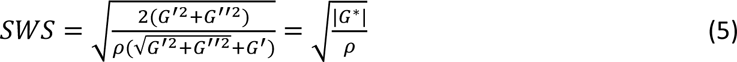

Typically, a mass density of *ρ* = 1000 kg/m^3^ is assumed in elastography. *SWS* is the parameter provided by THE without any further model assumptions. Therefore, we report *SWS* as a surrogate marker of stiffness without conversion to shear modulus or Young’s modulus in accordance with current guidelines for liver elastography (18). Diastolic *SWS* was automatically detected based on the recorded ECG and averaged within the diastolic phases. The correlation coefficient obtained from the displacement estimation was used to discard all diastolic frames with a correlation coefficient below 0.9, resulting in n=39±15 [13-114]) frames to be averaged. The final THE-SWS maps provided a surrogate marker of diastolic stiffness (see figure S1_5).

**Figure S1_5.**
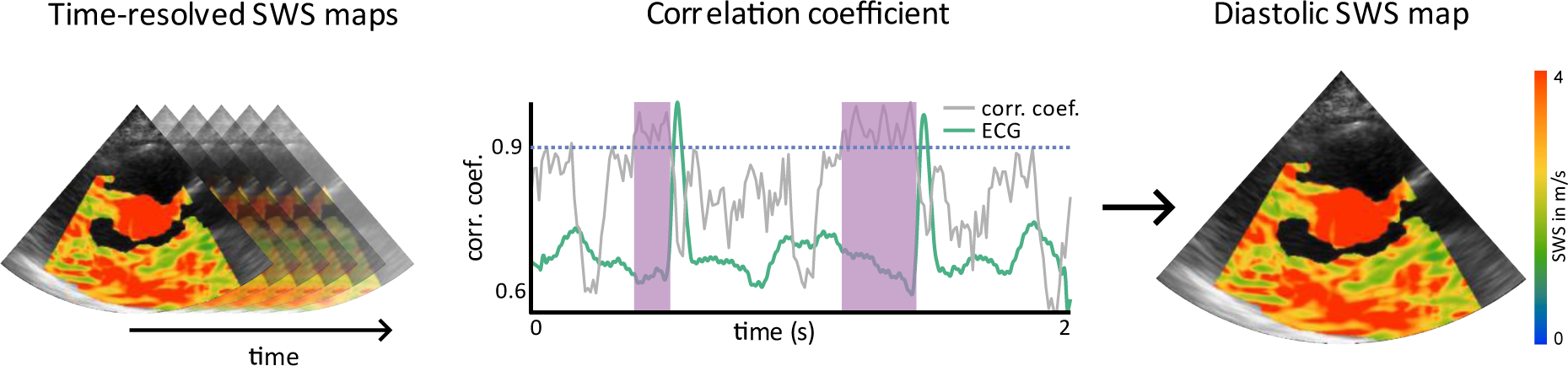
**Legend:** Averaging of diastolic SWS maps. Diastolic SWS maps were selected based on the recorded ECG and by applying a threshold of 0.9 to the correlation coefficient obtained from the displacement estimation. Selected frames were averaged to generate one SWS map that represents a surrogate marker for myocardial stiffness during the diastole.

It should be mentioned, that THE is currently limited to 2D and axial displacements whereas shear waves in the heart are 3D vector fields. Therefore, a geometric bias may occur when waves propagate obliquely through the image plane. However, the directional filters in the *k*-MDEV inversion support in-plane propagating waves while suppressing waves that cross the image plane obliquely. In addition, the cosine dependence of the geometric bias, as discussed in (19), minimizes the effect of oblique waves on the resulting *SWS* maps.

## References

1. McDonagh TA, Metra M, Adamo M et al. 2021 ESC Guidelines for the diagnosis and treatment of acute and chronic heart failure. Eur Heart J 2021;42:3599–3726.

2. Savarese G, Becher PM, Lund LH, Seferovic P, Rosano GMC, Coats A. Global burden of heart failure: A comprehensive and updated review of epidemiology. Cardiovasc Res 2022.

3. Paulus WJ, Tschöpe C, Sanderson JE et al. How to diagnose diastolic heart failure: a consensus statement on the diagnosis of heart failure with normal left ventricular ejection fraction by the Heart Failure and Echocardiography Associations of the European Society of Cardiology. Eur Heart J 2007;28:2539–50.

4. Obokata M, Reddy YNV, Borlaug BA. Diastolic Dysfunction and Heart Failure With Preserved Ejection Fraction: Understanding Mechanisms by Using Noninvasive Methods. JACC Cardiovasc Imaging 2020;13:245–257.

5. Nagueh SF, Smiseth OA, Appleton CP et al. Recommendations for the Evaluation of Left Ventricular Diastolic Function by Echocardiography: An Update from the American Society of Echocardiography and the European Association of Cardiovascular Imaging. J Am Soc Echocardiogr 2016;29:277–314.

6. Okada K, Kaga S, Abiko R et al. Novel echocardiographic method to assess left ventricular chamber stiffness and elevated end-diastolic pressure based on time-velocity integral measurements of pulmonary venous and transmitral flows. Eur Heart J Cardiovasc Imaging 2018;19:1260–1267.

7. Konofagou EE, D’hooge J, Ophir J. Myocardial elastography—a feasibility study in vivo. Ultrasound in Medicine & Biology 2002;28:475–482.

8. Villemain O, Correia M, Khraiche D et al. Myocardial Stiffness Assessment Using Shear Wave Imaging in Pediatric Hypertrophic Cardiomyopathy. JACC: Cardiovascular Imaging 2018;11:779–781.

9. Tzschatzsch H, Nguyen Trong M, Scheuermann T et al. Two-Dimensional Time-Harmonic Elastography of the Human Liver and Spleen. Ultrasound Med Biol 2016;42:2562–2571.

10. Dietrich CF, Bamber J, Berzigotti A et al. EFSUMB Guidelines and Recommendations on the Clinical Use of Liver Ultrasound Elastography, Update 2017 (Long Version). Ultraschall Med 2017;38:e16–e47.

11. Sack I. Magnetic resonance elastography from fundamental soft-tissue mechanics to diagnostic imaging. Nature Reviews Physics 2022:1–18.

12. Troelstra MA, Runge JH, Burnhope E et al. Shear wave cardiovascular MR elastography using intrinsic cardiac motion for transducer-free non-invasive evaluation of myocardial shear wave velocity. Scientific Reports 2021;11:1403.

13. Tzschatzsch H, Guo J, Dittmann F et al. Tomoelastography by multifrequency wave number recovery from time-harmonic propagating shear waves. Medical Image Analysis 2016;30:1–10.

14. Pernot M, Villemain O. Myocardial Stiffness Assessment by Ultrasound. JACC: Cardiovascular Imaging 2020;13:2314–2315.

15. Arani A, Arunachalam SP, Chang ICY et al. Cardiac MR elastography for quantitative assessment of elevated myocardial stiffness in cardiac amyloidosis. Journal of Magnetic Resonance Imaging 2017;46:1361–1367.

16. Cvijic M, Bézy S, Petrescu A et al. Interplay of cardiac remodelling and myocardial stiffness in hypertensive heart disease: a shear wave imaging study using high-frame rate echocardiography. European Heart Journal - Cardiovascular Imaging 2019;21:664–672.

17. Petrescu A, Santos P, Orlowska M et al. Velocities of Naturally Occurring Myocardial Shear Waves Increase With Age and in Cardiac Amyloidosis. JACC Cardiovasc Imaging 2019;12:2389–2398.

18. Santos P, Petrescu AM, Pedrosa J et al. Natural Shear Wave Imaging in the Human Heart: Normal Values, Feasibility, and Reproducibility. IEEE Transactions on Ultrasonics, Ferroelectrics, and Frequency Control 2019;66:442–452.

19. Pislaru C, Alashry MM, Ionescu F et al. Increased Myocardial Stiffness Detected by Intrinsic Cardiac Elastography in Patients With Amyloidosis: Impact on Outcomes. JACC Cardiovasc Imaging 2019;12:375–377.

20. Strachinaru M, Bosch JG, van Gils L et al. Naturally Occurring Shear Waves in Healthy Volunteers and Hypertrophic Cardiomyopathy Patients. Ultrasound in Medicine and Biology 2019;45:1977–1986.

21. Villemain O, Correia M, Mousseaux E et al. Myocardial Stiffness Evaluation Using Noninvasive Shear Wave Imaging in Healthy and Hypertrophic Cardiomyopathic Adults. JACC Cardiovasc Imaging 2019;12:1135–1145.

22. Song P, Bi X, Mellema DC et al. Quantitative Assessment of Left Ventricular Diastolic Stiffness Using Cardiac Shear Wave Elastography. Journal of Ultrasound in Medicine 2016;35:1419–1427.

23. Voigt J-U. Direct Stiffness Measurements by Echocardiography: Does the Search for the Holy Grail Come to an End?∗. JACC: Cardiovascular Imaging 2019;12:1146–1148.

24. Tadic M, Cuspidi C. Obesity and heart failure with preserved ejection fraction: a paradox or something else? Heart Fail Rev 2019;24:379–385.

25. Hudert CA, Tzschätzsch H, Guo J et al. US Time-Harmonic Elastography: Detection of Liver Fibrosis in Adolescents with Extreme Obesity with Nonalcoholic Fatty Liver Disease. Radiology 2018;288:99–106.

26. Kreft B, Tzschatzsch H, Schrank F et al. Time-Resolved Response of Cerebral Stiffness to Hypercapnia in Humans. Ultrasound Med Biol 2020;46:936–943.

27. Schaafs LA, Tzschätzsch H, Reshetnik A et al. Ultrasound Time-Harmonic Elastography of the Aorta: Effect of Age and Hypertension on Aortic Stiffness. Invest Radiol 2019;54:675–680.

28. Ruberg FL, Grogan M, Hanna M, Kelly JW, Maurer MS. Transthyretin Amyloid Cardiomyopathy: JACC State-of-the-Art Review. J Am Coll Cardiol 2019;73:2872–2891.

29. Maurer MS, Schwartz JH, Gundapaneni B et al. Tafamidis Treatment for Patients with Transthyretin Amyloid Cardiomyopathy. N Engl J Med 2018;379:1007–1016.

30. Garcia-Pavia P, Rapezzi C, Adler Y et al. Diagnosis and treatment of cardiac amyloidosis. A position statement of the European Society of Cardiology Working Group on Myocardial and Pericardial Diseases. Eur J Heart Fail 2021.

31. Hagendorff A, Fehske W, Flachskampf FA et al. Manual zur Indikation und Durchführung der Echokardiographie – Update 2020 der Deutschen Gesellschaft für Kardiologie. Der Kardiologe 2020;14:396–431.

32. Galderisi M, Cosyns B, Edvardsen T et al. Standardization of adult transthoracic echocardiography reporting in agreement with recent chamber quantification, diastolic function, and heart valve disease recommendations: an expert consensus document of the European Association of Cardiovascular Imaging. Eur Heart J Cardiovasc Imaging 2017;18:1301–1310.

33. Sun D, Schaff HV, Nishimura RA et al. Posterior Wall Thickness Associates With Survival Following Septal Myectomy for Obstructive Hypertrophic Cardiomyopathy. JACC: Heart Failure 2022;10:831–837.

34. Wassenaar PA, Eleswarpu CN, Schroeder SA et al. Measuring age-dependent myocardial stiffness across the cardiac cycle using MR elastography: A reproducibility study. Magn Reson Med 2016;75:1586–93.

35. Hieda M, Sarma S, Hearon CM, Jr. et al. One-Year Committed Exercise Training Reverses Abnormal Left Ventricular Myocardial Stiffness in Patients With Stage B Heart Failure With Preserved Ejection Fraction. Circulation 2021;144:934–946.

## Supporting Material References

1. Tzschatzsch H, Nguyen Trong M, Scheuermann T, Ipek-Ugay S, Fischer T, Schultz M, et al. Two-Dimensional Time-Harmonic Elastography of the Human Liver and Spleen. Ultrasound Med Biol. 2016;42(11):2562–71. PubMed PMID: 27567061.

2. Morr AS, Herthum H, Schrank F, Görner S, Anders MS, Lerchbaumer M, et al. Liquid-Liver Phantom: Mimicking the Viscoelastic Dispersion of Human Liver for Ultrasound- and MRI-Based Elastography. Invest Radiol. 2022;57(8):502–9. Epub 20220223. doi: 10.1097/rli.0000000000000862. PubMed PMID: 35195086.

3. Heucke N, Wuensch T, Mohr J, Kaffarnik M, Arsenic R, Sinn B, et al. Non-invasive structure-function assessment of the liver by 2D time-harmonic elastography and the dynamic Liver MAximum capacity (LiMAx) test. J Gastroenterol Hepatol. 2019;34(9):1611–9. Epub 2019/02/14. doi: 10.1111/jgh.14629. PubMed PMID: 30756433.

4. Marticorena Garcia SR, Grossmann M, Lang ST, Nguyen Trong M, Schultz M, Guo J, et al. Full-Field-of-View Time-Harmonic Elastography of the Native Kidney. Ultrasound Med Biol. 2018;44(5):949–54. PubMed PMID: 29478787.

5. Kreft B, Tzschatzsch H, Schrank F, Bergs J, Streitberger KJ, Waldchen S, et al. Time-Resolved Response of Cerebral Stiffness to Hypercapnia in Humans. Ultrasound Med Biol. 2020;46(4):936–43. Epub 2020/02/01. doi: 10.1016/j.ultrasmedbio.2019.12.019. PubMed PMID: 32001088.

6. Schaafs LA, Tzschätzsch H, Reshetnik A, van der Giet M, Braun J, Hamm B, et al. Ultrasound Time-Harmonic Elastography of the Aorta: Effect of Age and Hypertension on Aortic Stiffness. Invest Radiol. 2019;54(11):675–80. doi: 10.1097/rli.0000000000000590. PubMed PMID: 31299035.

7. Ormachea J, Castaneda B, Parker KJ. Shear Wave Speed Estimation Using Reverberant Shear Wave Fields: Implementation and Feasibility Studies. Ultrasound in Medicine and Biology. 2018;44(5):963–77. doi: 10.1016/j.ultrasmedbio.2018.01.011.

8. Parker KJ, Huang SR, Musulin RA, Lerner RM. Tissue response to mechanical vibrations for “sonoelasticity imaging”. Ultrasound Med Biol. 1990;16(3):241–6. doi: 10.1016/0301-5629(90)90003-u. PubMed PMID: 2194336.

9. Civale J, Parasaram V, Bamber JC, Harris EJ. High frequency ultrasound vibrational shear wave elastography for preclinical research. Physics in Medicine & Biology. 2022;67(24):245005. doi: 10.1088/1361-6560/aca4b8.

10. Hudert CA, Tzschätzsch H, Guo J, Rudolph B, Bläker H, Loddenkemper C, et al. US Time-Harmonic Elastography: Detection of Liver Fibrosis in Adolescents with Extreme Obesity with Nonalcoholic Fatty Liver Disease. Radiology. 2018;288(1):99–106. Epub 20180515. doi: 10.1148/radiol.2018172928. PubMed PMID: 29762096.

11. Tzschatzsch H, Kreft B, Schrank F, Bergs J, Braun J, Sack I. In vivo time-harmonic ultrasound elastography of the human brain detects acute cerebral stiffness changes induced by intracranial pressure variations. Sci Rep. 2018;8(1):17888. Epub 2018/12/19. doi: 10.1038/s41598-018-36191-9. PubMed PMID: 30559367; PubMed Central PMCID: PMC6297160.

12. Kasai C, Namekawa K, Koyano A, Omoto R. Real-Time Two-Dimensional Blood Flow Imaging Using an Autocorrelation Technique. IEEE Transactions on Sonics and Ultrasonics. 1985;32(3):458–64. doi: 10.1109/T-SU.1985.31615.

13. Meyer T, Kreft B, Bergs J, Antes E, Anders MS, Wellge B, et al. Stiffness pulsation of the human brain detected by non-invasive time-harmonic elastography. Frontiers in Bioengineering and Biotechnology. 2023;11. doi: 10.3389/fbioe.2023.1140734.

14. Tzschatzsch H, Guo J, Dittmann F, Hirsch S, Barnhill E, Johrens K, et al. Tomoelastography by multifrequency wave number recovery from time-harmonic propagating shear waves. Medical Image Analysis. 2016;30:1–10. doi: 10.1016/j.media.2016.01.001. PubMed PMID: WOS:000373546800001.

15. Herthum H, Shahryari M, Tzschatzsch H, Schrank F, Warmuth C, Gorner S, et al. Real-Time Multifrequency MR Elastography of the Human Brain Reveals Rapid Changes in Viscoelasticity in Response to the Valsalva Maneuver. Front Bioeng Biotechnol. 2021;9(335):666456. Epub 2021/05/25. doi: 10.3389/fbioe.2021.666456. PubMed PMID: 34026743; PubMed Central PMCID: PMC8131519.

16. Manduca A, Lake DS, Kruse SA, Ehman RL. Spatio-temporal directional filtering for improved inversion of MR elastography images. Med Image Anal. 2003;7(4):465–73. doi: 10.1016/s1361-8415(03)00038-0. PubMed PMID: 14561551.

17. Sack I. Magnetic resonance elastography from fundamental soft-tissue mechanics to diagnostic imaging. Nature Reviews Physics. 2022:1–18. doi: 10.1038/s42254-022-00543-2.

18. Dietrich CF, Bamber J, Berzigotti A, Bota S, Cantisani V, Castera L, et al. EFSUMB Guidelines and Recommendations on the Clinical Use of Liver Ultrasound Elastography, Update 2017 (Long Version). Ultraschall Med. 2017;38(4):e16–e47. Epub 20170413. doi: 10.1055/s-0043-103952. PubMed PMID: 28407655.

19. Tzschätzsch H, Ipek-Ugay S, Guo J, Streitberger KJ, Gentz E, Fischer T, et al. In vivo time-harmonic multifrequency elastography of the human liver. Phys Med Biol. 2014;59(7):1641–54. Epub 20140310. doi: 10.1088/0031-9155/59/7/1641. PubMed PMID: 24614751.

